# Recognition of Category A bioterrorism agent syndromes by final-year medical students in the UK: a pilot study

**DOI:** 10.64898/2026.08.21.26361035

**Authors:** Richard C Armitage, Charlotte Hammer

**Affiliations:** University of Cambridge, Centre for the Study of Existential Risk; University of Nottingham, School of Medicine

## Abstract

**Background:** Early recognition of presentations consistent with the deliberate release of a Category A bioterrorism agent is essential for rapid isolation, public health notification, and containment. The ability of UK clinicians-in-training to recognise these syndromes is unstudied. This pilot assessed final-year UK medical students’ ability to recognise these syndromes.

**Methods:** A pilot cross-sectional online survey of final-year UK medical students used single-best-answer clinical vignettes depicting syndromes associated with Category A bioterrorism agents (BT vignettes) and clinically overlapping non-bioterrorism syndromes (NBT vignettes). Performance was summarised as the proportion of vignettes correctly identified, with primary analysis comparing within-participant BT and NBT performance.

**Results:** Twenty-five participants completed the survey. Participants performed worse on BT vignettes (M = 0.55) than on NBT vignettes (M = 0.81), with a within-participant difference of −0.26 (95% CI [-0.35, −0.18]; t(24) = -6.33, p < 0.001; Cohen’s dz = -1.27). Botulism (96.0%) and Ebola virus disease (88.0%) were recognised by most participants, while anthrax (40.0%), pneumonic plague (28.0%), and smallpox (24.0%) were recognised by fewer than half.

**Conclusion:** This pilot provides the first UK evidence of a substantial diagnostic deficit in final-year medical students’ recognition of Category A bioterrorism agent syndromes.

## INTRODUCTION

This study examines the ability of UK final-year medical students to recognise the syndromes of Category A bioterrorism agents – pathogens designated as posing the greatest public health risk in the event of deliberate release – addressing a significant gap in the UK evidence base on clinical preparedness for bioterrorism.

The weaponisation of contagious disease dates from antiquity;^1,2^ pre-modern examples are summarised in Table 1.^2,3^ The modern era began with the late-19th-century foundation of microbiology, which enabled the isolation, production, and dissemination of specific pathogens at scale. Clandestine First World War programmes – notably Germany’s contamination of enemy animal feed with Bacillus anthracis and Burkholderia mallei^4,5^ – prompted the 1925 Geneva Protocol, which outlawed the use of biological weapons.^2,3^ Major state programmes followed in the US, UK, France, the Soviet Union, and others.^2,3,6,7^ The 1975 BWC subsequently prohibited possession except for peaceful purposes and mandated destruction of stockpiles^8,9^ but has not eliminated state-level research; Iraq’s Gulf War-era programme^2^ and the 1979 Sverdlovsk accidental anthrax release^3,10,11^ illustrate this persistence.

**Table 1:** Examples of biological warfare in the pre-microbiological era.

| Year | Event |
| --- | --- |
| 14th century BC | Rams infected with tularaemia are sent by The Hittites to their enemies |
| 4th century BC | Scythian archers dip their arrows into decomposing adder cadavers and human blood |
| 1155 | Water wells in Tortona (Italy) are poisoned with human bodies by Emperor Barbarossa |
| 1346 | Bodies of plague victims are catapulted over the city walls of Caffa by Mongols |
| 1495 | In Naples (Italy), Spanish sell wine mixed with blood of leprosy patients to their French adversaries |
| 1650 | Saliva from rabid dogs is fired by Polish army towards their enemies |
| 1710 | In Reval (Estonia), plague cadavers are hurled onto Swedish troops by Russian army |
| 1763 | British officers provide Native Americans with blankets from smallpox hospitals |
| 1797 | To increase the transmission of malaria among their enemy, Napoleonic armies flood the plains around Mantua (Italy) |
| 1863 | During the American Civil War, Confederates sell clothing from yellow fever and smallpox patients to Union troops |

A principal contemporary concern is bioterrorism – the use of biological weapons by non-state actors.^2^ Notable incidents include the 1984 Rajneesh cult contamination of Oregon salad bars with Salmonella typhimurium;^12^ the Aum Shinrikyo cult’s biological weapons programme alongside its 1995 Tokyo sarin attack;^3,12^ and the 2001 US ’anthrax letters’, which caused 22 infections and five deaths and demonstrated bioterrorism’s capacity for disproportionate psychological and political impact.^2,13^

The CDC classifies bioweapons-capable agents into three tiers, with Category A representing the highest priority.^14,15^ Category A agents are readily disseminated or transmitted between individuals, cause substantial mortality, provoke widespread public alarm, and require specialised public health preparedness.^16,17^ They typically share at least three of five features: low infective dose with high contagiousness; difficulty in recognition as a deliberate attack; amenability to mass production, storage, and weaponisation; limited pre-existing immunity or treatment; and feasibility of perpetrator self-protection.^18^

The six Category A agents are anthrax (Bacillus anthracis), botulism (Clostridium botulinum toxin), plague (Yersinia pestis), smallpox (Variola major), tularaemia (Francisella tularensis), and viral haemorrhagic fevers (filoviruses such as Ebola and Marburg, and arenaviruses such as Lassa and Machupo).^17,19,20^ Their modes of transmission and clinical features are summarised in Table 2.^14,21–26^ Most can be aerosolised and disseminated across wide areas, with further routes including postal systems and intentional introduction of infected vectors.^25,27^ Their clinical manifestations largely overlap with far more common illnesses, particularly early on, and most occur naturally albeit rarely, so bioterrorist attacks are difficult to distinguish from natural outbreaks.^16,28^ Even so, a single confirmed case of smallpox, of anthrax without occupational risk factors, of viral haemorrhagic fever without relevant travel history, or of plague or tularaemia without ecological exposure should raise strong suspicion of deliberate release.^27,29^

**Table 2:** Summary of the clinical features of Category A bioterrorism agents.

| Category A bioterrorism agent | Modes of transmission | Clinical features |
| --- | --- | --- |
| Anthrax | Inhalation<br>Cutaneous (much lower mortality) | Non-specific prodrome of fever, dyspnoea, cough, and chest discomfort following exposure to infectious spores<br>After 2-4 days, patients typically deteriorate rapidly into respiratory failure and haemodynamic collapse<br>Chest radiograph may demonstrate thoracic oedema and mediastinal widening |
| Botulism | Ingestion<br>Inhalation | Symmetrical cranial neuropathies (such as ptosis, dysphagia, and dysarthria)<br>Visual disturbances (such as blurred vision or diplopia)<br>Symmetrical descending weakness progresses in a proximal to distal pattern<br>Respiratory dysfunction may occur due to respiratory muscle paralysis or upper airway obstruction<br>Sensory function remains intact |
| Plague | Vector-borne<br>Contact with bodily fluids<br>Respiratory droplet inhalation | Fever, cough producing mucopurulent sputum, haemoptysis, and chest pain<br>Chest radiograph typically demonstrates bronchopneumonia |
| Smallpox | Respiratory droplet inhalation<br>Direct contact with sore | Non-specific prodrome of fever and myalgia for 2-4 days prior to rash onset, resembling other acute viral illnesses such as influenza<br>Vesicular/pustular rash is typically most prominent on the face and extremities |
|  |  | • Lesions develop synchronously, appearing at the same stage of evolution<br>• Differs from *Varicella zoster* virus (which causes chickenpox), in which the rash is most prominent on the trunk and lesions develop in successive crops over several days, resulting in lesions at various stages of development and resolution |
| Tularaemia | • Vector-borne<br>• Contact with infected animals<br>• Contaminated food or water<br>• Inhalation | • Abrupt onset of an acute, non-specific febrile illness 3-5 days after exposure<br>• Pleuropneumonitis develops over subsequent days |
| Viral haemorrhagic fevers |  | • Abrupt onset fever, myalgia, and headache<br>• Nausea and vomiting, abdominal pain, diarrhoea, chest pain, cough, and pharyngitis<br>• A maculopapular rash, predominantly on the trunk<br>• Haemorrhagic manifestations such as petechiae, ecchymoses, and frank haemorrhage |

Recent developments in synthetic biology and AI are reshaping the bioweapons threat.^30^ Benchtop DNA synthesis devices and the broader democratisation of synthesis capacity^31^ can partially bypass provider-side safeguards such as sequence screening,^32^ raising concern that malign actors might more easily obtain genetic material relevant to high-consequence pathogens, including Category A agents.^33–35^ Convergence between AI and the life sciences ("AIxBio") may simultaneously lower the floor for moderately skilled actors and raise the ceiling of harm by sophisticated ones.^30,36–41^ These evolving capabilities heighten the importance of frontline physicians’ ability to recognise Category A bioterrorism agent syndromes.

Early recognition of a bioterrorism attack is critical for rapid isolation of affected individuals,^16^ containment of communicable pathogens,^27^ and minimisation of morbidity and mortality.^14,42^ Patients exposed to deliberately released agents are likely to present initially to Emergency Departments, Acute Medicine Receiving Units, and other frontline services,^43^ so timely recognition enables clinical management, public health notification, and rapid containment,^43–45^ particularly for those practising in emergency medicine, primary care, general internal medicine, infectious diseases, microbiology, virology, and dermatology.^46,47^ Although bioterrorism syndromes may closely resemble naturally occurring outbreaks,^16^ subtle but critical clinical differences can enable their recognition.^28^

The Health Protection (Notification) Regulations 2010 require UK registered medical practitioners to notify suspected or confirmed cases of specified notifiable diseases – including anthrax, botulism, plague, smallpox, and viral haemorrhagic fever^48,49^ – placing a statutory duty on physicians to recognise these conditions. The International Health Regulations (2005), to which the UK is a signatory, similarly require notification to the WHO of deliberate biological releases.^50^ Plague and viral haemorrhagic fevers are included within the UK HCID framework, with cases to be rapidly transferred to a designated HCID Treatment Centre.^51^

Public Health England’s 2018 guidance emphasises that early recognition of a covert biological release depends on physicians remaining aware of the possibility and willing to alert their microbiologist, infectious disease clinician, and Health Protection Team on suspicion.^52^ NHS England^53^ and the UK Biological Security Strategy^54^ similarly highlight this need. The General Medical Council requires physicians to adequately assess a patient’s condition including symptoms,^55^ and newly qualified physicians to propose a prioritised differential diagnosis^56^ – requirements directly applicable to the recognition of Category A bioterrorism syndromes.

Despite this clinical and regulatory importance, physicians’ diagnostic capability for Category A agents remains poorly understood. Earlier studies have largely relied on self-reported confidence ^57,58^ or broad knowledge measures rather than tested syndrome recognition.^59,60^ The most robust assessment to date is the 2003-2004 work of Cosgrove et al., which used 16 vignettes; correct diagnoses of Category A diseases were provided in only 46.8% of cases on average, rising to 79.0% after a brief didactic module.^61^ No comparable work appears to have been conducted in the UK, and the two-decade interval since Cosgrove et al. – during which the threat landscape has evolved substantially – further motivates contemporary assessment.

Evidence on curricular coverage is also limited. In the US, only 53% of emergency medicine residency programmes included formal bioterrorism training in 1999,^62^ and the emerging field of terror medicine has highlighted the absence of structured pre-qualification content.^63–67^ In the UK and Ireland, only 17.65% of medical schools included specific teaching on biological weapons and bioterrorism in 2012,^68^ with no further UK or US assessment since.

A definitive study addressing this UK evidence gap would require distribution through the MSC, now the conventional and increasingly required route through which UK medical schools accept research surveys.^69^ MSC approval is lengthy and exceeded the time available; a pilot was therefore undertaken to provide preliminary findings, test the feasibility of the instrument and recruitment approach, and inform a subsequent definitive study.

The aim was to generate preliminary evidence on the ability of final-year UK medical students – the pre- qualification cohort closest to independent NHS practice – to recognise the syndromes of Category A bioterrorism agents using validated clinical vignettes. Secondary objectives were to assess variation across individual agents; the feasibility of comparing undergraduate and graduate-entry programmes; and the feasibility of the instrument and recruitment approach.

The aim was to generate preliminary evidence on the ability of final-year UK medical students to recognise the syndromes of Category A bioterrorism agents using validated clinical vignettes. Final-year students are not themselves the frontline – practising clinicians in acute specialties are the population on whose recognition detection ultimately depends – but they are a sensible place to start. Newly-qualified doctors join the frontline workforce immediately, so the level of recognition students have reached by qualification is important; it also gives a baseline for the study of practising clinicians that should follow. Secondary objectives were to assess variation across individual agents; the feasibility of comparing undergraduate and graduate-entry programmes; and the feasibility of the instrument and recruitment approach.

## METHODS

The study was a pilot cross-sectional online survey of final-year UK medical students. Online deployment enabled standardised delivery and efficient data collection across multiple medical schools. The study population was final-year medical students at UK medical schools, including both undergraduate and graduate-entry programmes. UK medical schools with a final-year cohort in 2025- 2026 are listed in Table 3;^70^ recently established schools without a final-year cohort are excluded.

**Table 3:**
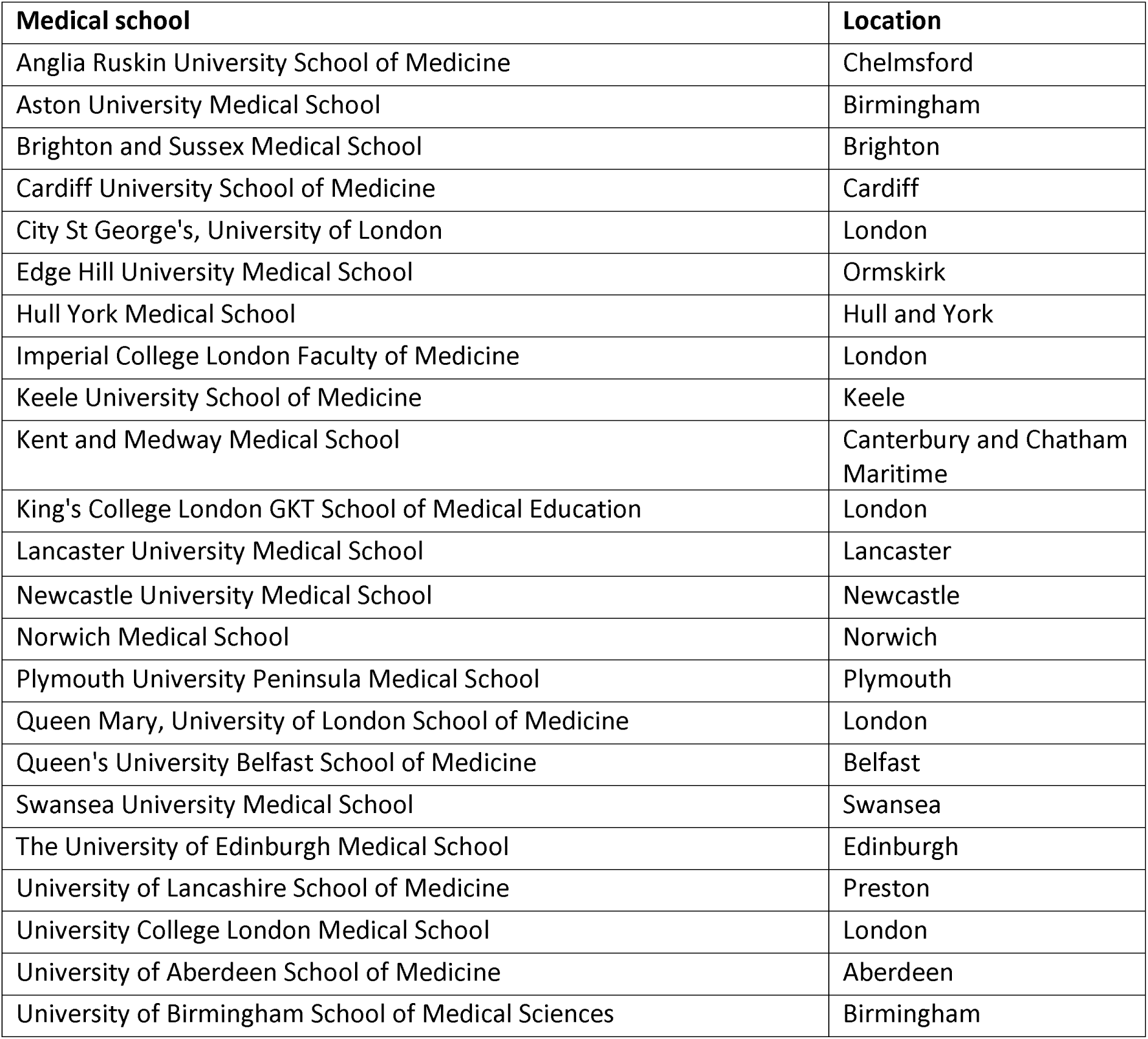
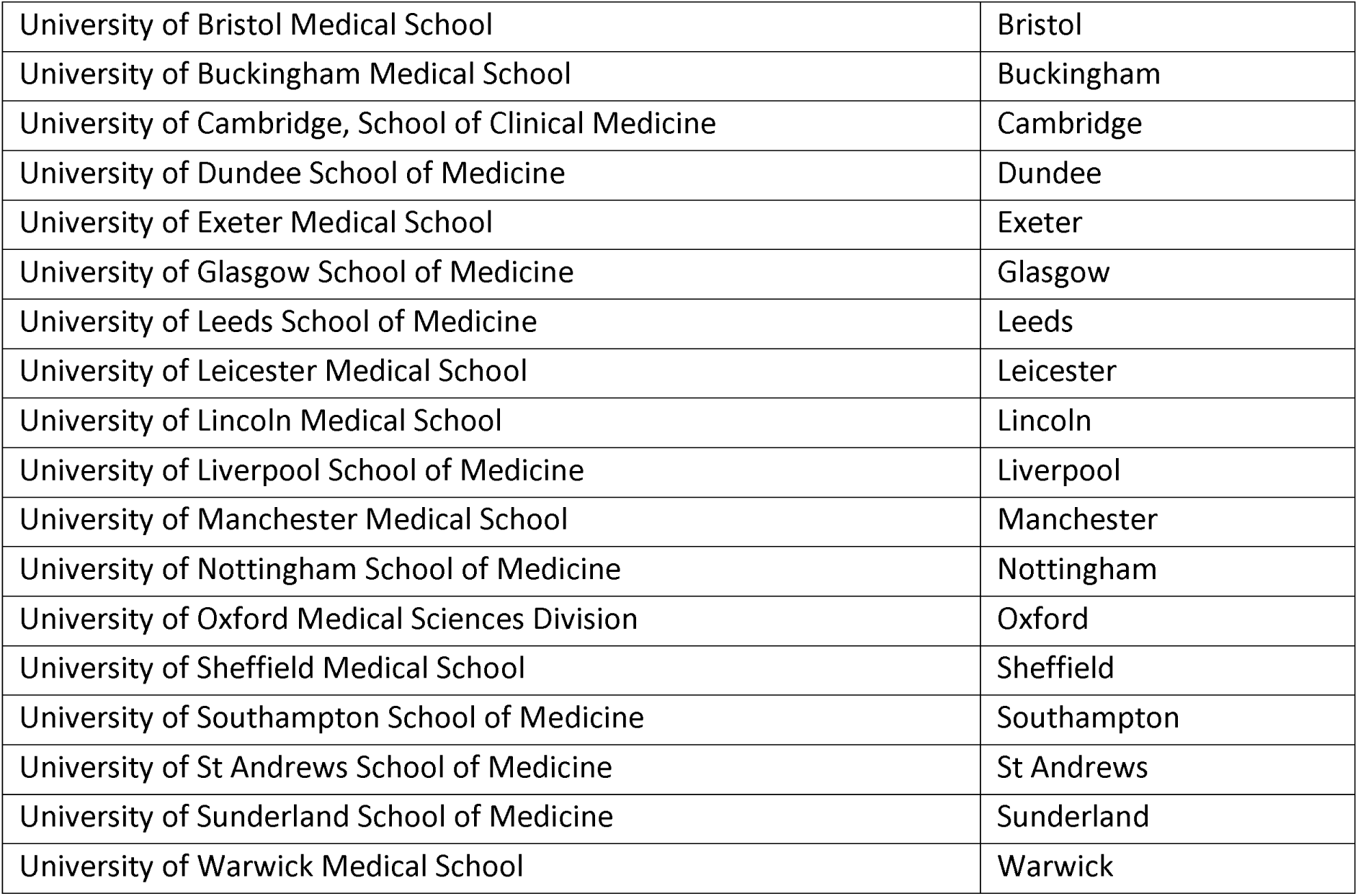
UK medical schools with final-year medical students in the 2025-2026 academic year.

In the UK, dissemination of research surveys to medical students is now conventionally conducted by the Education Leads Advisory Group of the MSC,^69^ with some schools accepting surveys only through this route. As the MSC process exceeded the time available, the survey was distributed directly to Heads of School at each eligible UK medical school. Each Head was contacted once via publicly available institutional email from the first author’s University of Cambridge account, asked to distribute the survey invitation by email to their final-year students, and asked to reply with the cohort size to enable response rate calculation; no repeat invitations were issued. The Participant Information Sheet described the survey as presenting brief clinical vignettes with multiple-choice answers, with no reference to the bioterrorism focus, to minimise priming bias. Prior to participation, participants confirmed agreement with the Participant Consent Form. A self-selected volunteer sampling strategy was used, with inclusion determined by voluntary response to the survey invitation. The implications of this non-probability approach for generalisability are addressed in the Discussion.

Data were collected via an online survey on the KoboToolbox platform, comprising a demographic component and a clinical component. The only demographic variable collected was programme type (undergraduate or graduate-entry). Email addresses were collected solely to identify and exclude duplicate submissions, and were deleted after the survey closed; email domains were inspected during deduplication and used to characterise the institutional spread of the sample.

The clinical component consisted of 18 single-best-answer multiple-choice questions, each presenting a brief clinical vignette with five response options, with the position of the correct answer varied across vignettes. Five vignettes depicted Category A bioterrorism syndromes as defined by the CDC – anthrax, botulism, plague, smallpox, and Ebola virus disease (the BT vignettes). The remaining 13 served as distractors describing clinical syndromes unrelated to bioterrorism agents (the NBT vignettes); these share substantial clinical overlap with Category A agents but occur much more commonly, reducing response bias and concealing the study’s focus. No tularaemia vignette was developed because its syndrome cannot be reliably distinguished from naturally occurring infections – particularly influenza and atypical pneumonias – on clinical presentation alone.^25,61^

Content validity was established through iterative expert review by three consultant physicians with specialist expertise in microbiology, virology, and infectious diseases, with each vignette revised until consensus was reached on clinical accuracy and on the substantial clinical overlap of the distractors. The vignettes and response options are presented in Table 4.

**Table 4:** Clinical vignettes and diagnosis options used in the survey instrument.

| Number | Question | Options |
| --- | --- | --- |
| 1 | A patient with rapid onset fever, chills, myalgia, non-productive cough, sore throat, fatigue and headache. | Diphtheria<br>Norovirus<br>Epstein-Barr virus<br>Strep throat<br>Influenza* |
| 2 | An afebrile patient with acute chest pain, shortness of breath and haemoptysis. They recently travelled on a 12-hour flight. | Myocardial infarction<br>*Strep pneumoniae*<br>Anthrax<br>Pulmonary embolism*<br>Tuberculosis |
| 3 | A hospitalised patient who recently completed a course of antibiotics develops acute profuse watery and bloody diarrhoea, fever and abdominal pains. | Cholera<br>Salmonellosis<br>*Clostridioides difficile* *<br>Giardiasis<br>Norovirus |
| 4 | A febrile patient with a widespread rash that occurred over 24 hours, now has widespread firm, deep-seated pustules in the same stage of development. | Meningococcal sepsis<br>Chickenpox<br>Molluscum contagiosum<br>Smallpox<sup>†</sup><br>Coxsackie virus |
| 5 | An oncology patient with gradually worsening dry cough, breathlessness and decreasing exercise tolerance on a background of fever, weight loss, and sweats. | Influenza<br>Tuberculosis<br>Pneumocystis pneumonia*<br>*Strep pneumoniae*<br>Pertussis |
| 6 | A febrile patient with bilateral parotitis following a prodrome of fever headache, malaise, myalgia and anorexia. | Lyme disease<br>Mumps*<br>Typhoid<br>Measles<br>Tuberculosis |
| 7 | A patient with 5 days of fever, fatigue, and muscle, joint and abdominal pains, then develops diarrhoea, vomiting, and unexplained bleeding and bruising. | *Strep pneumoniae*<br>Pertussis<br>Ebola virus disease<sup>†</sup><br>Influenza<br>Pulmonary embolism |
| 8 | A patient with ascending weakness starting in the lower extremities 1 week after an episode of diarrhoea and vomiting. | Botulism<br>Guillain-Barre syndrome*<br>Eaton-Lambert syndrome<br>Myasthenia gravis<br>Lyme disease |
| 9 | A febrile patient with vesicular rash in different stages of development spread over the torso and extremities. | Coxsackie virus<br>Smallpox virus<br>Meningococcal sepsis<br>Varicella virus*<br>*PVL-Staph aureus* |
| 10 | A febrile patient with fatigue, cough, haemoptysis, chest pains, shortness of breath, nausea, vomiting and headache. Chest examination demonstrates abnormal lung sounds and a pleural effusion. Chest x-ray shows a significantly widened mediastinum. | Pulmonary embolism<br>Influenza<br>*Strep pneumoniae*<br>Anthrax<sup>†</sup><br>Pertussis |
| 11 | A febrile patient with headache, neck stiffness, photophobia and confusion. They develop a purpuric rash. | Encephalitis<br>Influenza<br>Viral meningitis<br>Guillain-Barre syndrome<br>Meningococcal sepsis* |
| 12 | A patient who moved to the UK from India 2 years ago presents with 6 months of weight loss, fevers, night sweats, worsening shortness of breath, productive cough and haemoptysis. | Ebola virus disease<br>*Strep pneumoniae*<br>HIV/AIDS<br>Tuberculosis*<br>Malaria |
| 13 | A 29-year-old woman with 2 days of cranial nerve abnormalities and descending paralysis with autonomic dysfunction. She ate some tinned food 3 days ago. | Myasthenia gravis<br>Guillain-Barre syndrome<br>Botulism<sup>†</sup><br>Eaton-Lambert syndrome<br>Lyme disease |
| 14 | A patient who recently returned from Sub-Saharan Africa with high fever, chills, rigors, headache, myalgia, cough, gastrointestinal upset, jaundice, splenomegaly and hepatomegaly. | Dengue fever<br>Zika virus<br>Chikungunya virus<br>Yellow fever<br>Malaria* |
| 15 | A febrile delirious patient with cough, blood-tinged sputum, and tachypnoea. Chest x-ray shows consolidation in the left lower lobe. | Influenza<br>Epstein-Barr virus<br>Pneumonic plague |
|  |  | Tuberculosis<br>*Strep pneumoniae* * |
| 16 | A healthcare worker presents with recurrent skin boils over numerous months. The patient is afebrile and otherwise well. | Mumps<br>HIV/AIDS<br>PVL- *Staph aureus* *<br>Herpes simplex<br>Varicella zoster |
| 17 | A febrile 18-year-old patient with cervical lymphadenopathy, sore throat, palatal petechiae and splenomegaly. The sore throat gets worse after several days. | Mumps<br>Epstein-Barr virus*<br>Measles<br>Strep throat<br>Pertussis |
| 18 | A febrile patient with haemoptysis, tachycardia, tachypnoea, and right middle lobe infiltrate. They were exposed to a family member who died of pneumonia 4 days earlier. | *Strep pneumoniae*<br>Brucellosis<br>Influenza<br>Tuberculosis<br>Pneumonic plague<sup>†</sup> |

The primary outcome measure was the BT proportion (BT vignettes correctly identified divided by 5). Secondary outcomes were the proportion correctly identifying each individual Category A agent; the number of participants enrolled on undergraduate and graduate-entry programmes; and indicators of feasibility (number of disseminating schools, overall response rate, and data completeness).

Analyses used R version 4.5.2 with significance at p < 0.05; a complete-case analysis was used. It was hypothesised that participants would perform less well on BT than on NBT vignettes. The BT proportion, the NBT proportion (correct NBT vignettes divided by 13), and the within-participant difference (BT minus NBT) were summarised using mean, SD, median, IQR, and range. The primary analysis was a paired-samples t-test of BT and NBT proportions, benchmarking each participant’s BT performance against their own NBT performance to control for individual-level variation; normality was assessed visually and with the Shapiro-Wilk test, Cohen’s d_z_ was reported as the effect size, and a Wilcoxon signed-rank test was conducted as sensitivity. Planned secondary analyses comprised per-agent recognition with 95% Wilson confidence intervals; a programme-type comparison via independent-samples t-test (or Mann-Whitney U if normality was violated), contingent on sufficient recruitment; and descriptive characterisation of feasibility.

Ethical approval was provided by the University of Cambridge’s Institute of Technology and Humanity Research Ethics Committee (ITH-REC reference: 25.3) on 09 December 2025.

## RESULTS

The survey was open between 16 February 2026 and 11 May 2026 (85 days). Of the 41 UK medical schools with a final-year cohort that were invited, seven (17.1%) declined to disseminate the survey (predominantly citing the absence of MSC approval), six (14.6%) confirmed dissemination, and 28 (68.3%) did not reply. Cohort sizes were supplied by four of the six confirming schools (range 130-303 students, total 805). A total of 25 participants completed the survey, of whom 24 (96.0%) were enrolled on undergraduate programmes and one (4.0%) on a graduate-entry programme; the planned subgroup analysis was therefore not feasible. There were no missing data.

A definitive response rate could not be calculated because cohort sizes were unavailable for two confirming schools and dissemination by non-responding schools could not be excluded; using the 805 students from the four schools with known cohort sizes as a minimum denominator generates an upper-bound response rate of 3.1% (25/805), with the true rate almost certainly lower. Institutional email domains identified at least five distinct schools represented in the sample, all corresponding to confirming schools; additional schools may be represented among participants who used personal email addresses.

The mean overall score across all 18 vignettes was 0.74 (SD = 0.13), indicating that participants correctly recognised the syndrome described in approximately three-quarters of vignettes on average.

Participants performed substantially worse on BT vignettes (M = 0.55, SD = 0.23, median = 0.60, IQR = 0.40-0.80, range 0.20-1.00) than on NBT vignettes (M = 0.81, SD = 0.11, median = 0.85, IQR = 0.77-0.92, range 0.54-1.00) (see Figure 1). Nearly half of participants (12 of 25, 48.0%) correctly recognised two or fewer of the five Category A syndromes (BT score ≤0.40). BT performance was also markedly more variable: the BT IQR (0.40) was more than two and a half times that of the NBT (0.15), reflecting heterogeneous preparation across participants.

**Figure 1:**
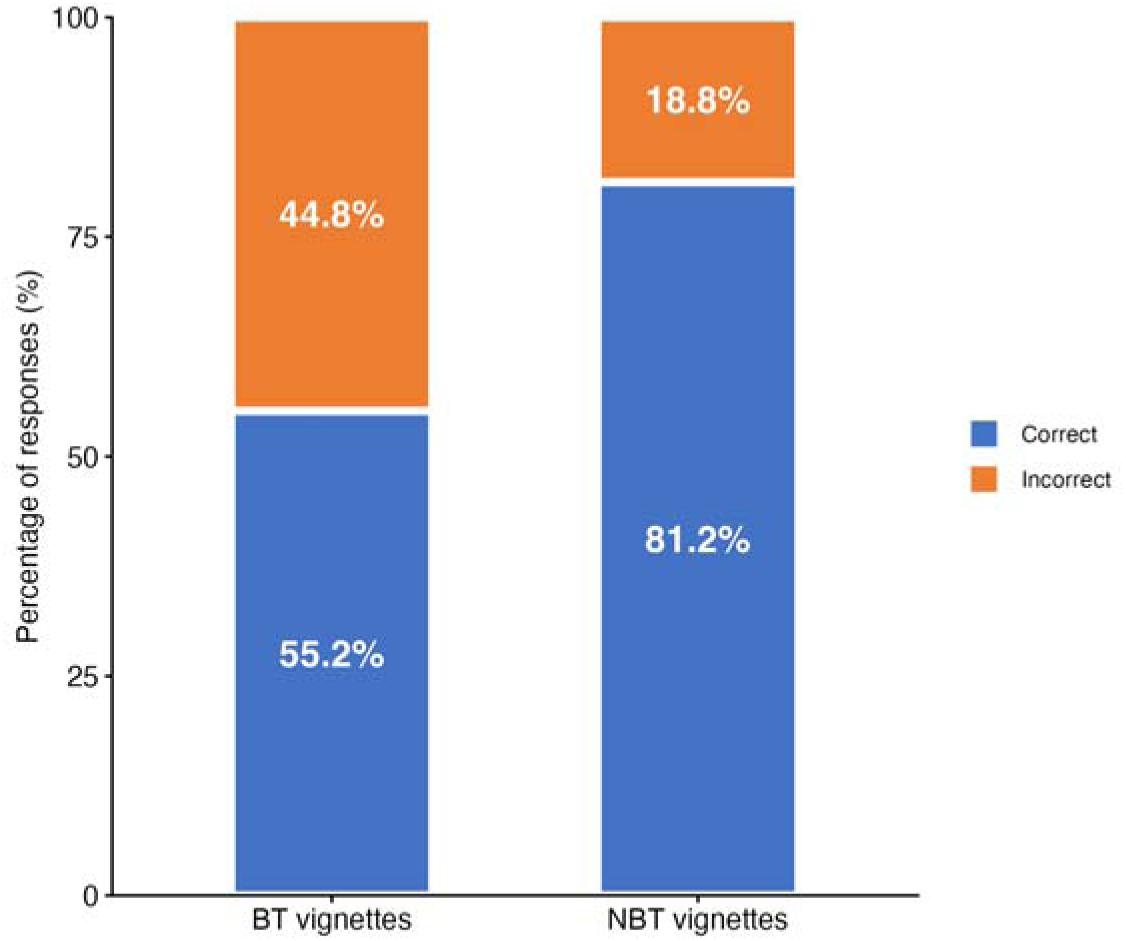
Percentage of responses correctly identifying the syndrome described in BT and NBT vignettes (n = 25 participants × 5 BT vignettes = 125 BT responses; n = 25 participants × 13 NBT vignettes = 325 NBT responses).

The mean within-participant difference was −0.26 (SD = 0.21, median = −0.22, IQR = −0.45 to −0.12, range −0.72 to +0.03). The distribution did not deviate significantly from normality (Shapiro-Wilk W = 0.95, p = 0.19). The paired-samples t-test yielded a highly significant mean difference of −0.26 (95% CI [-0.35, - 0.18]; t(24) = -6.33, p < 0.001), indicating that on average participants scored approximately 26 percentage points lower on BT vignettes than on NBT vignettes; Cohen’s d_z_ was -1.27, substantially exceeding the conventional threshold for a large effect (0.8). A Wilcoxon signed-rank test gave a concordant result (V = 3, p < 0.001; matched-pairs rank-biserial correlation r = 0.98). The effect was highly consistent: 24 of the 25 participants scored lower on BT than on NBT vignettes, with the remaining participant showing approximately equal performance (difference = +0.03); no participant scored meaningfully higher on BT than NBT vignettes (see Figure 2 and Table 5).

**Figure 2:**
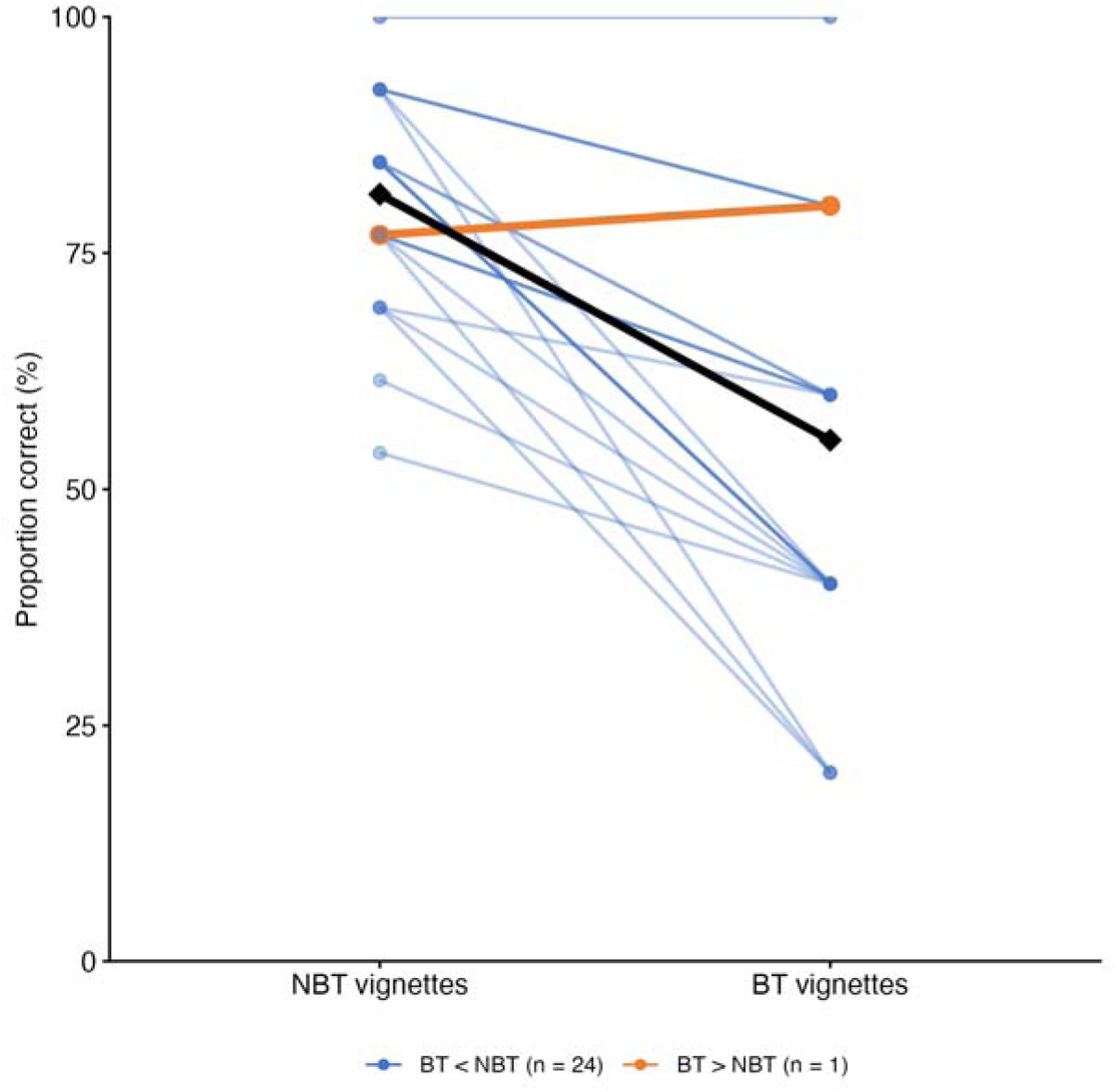
Within-participant comparison of recognition of BT and NBT vignettes (n = 25). Each thin line represents one participant: blue lines indicate participants whose BT performance was below their NBT performance (n = 24); the orange line indicates the participant whose BT performance exceeded their NBT performance (n = 1). The group mean is shown as a thick black line with diamond markers.

**Table 5:** Descriptive statistics for performance on the BT and NBT groups of vignettes (n = 25). Note: All values for BT and NBT groups are proportions correct. Paired-samples t-test (t(24) = -6.33, p < 0.001); 95% CI for the mean difference [-0.35, −0.18]; Cohen’s d_z_ = -1.27. Wilcoxon signed-rank test (sensitivity analysis): V = 3, p < 0.001; matched-pairs rank-biserial correlation r = 0.98. Shapiro-Wilk test of the within-participant differences: W = 0.95, p = 0.19.

| Group | Vignettes | Mean | *SD* | Median | IQR | Range |
| --- | --- | --- | --- | --- | --- | --- |
| BT (Category A bioterrorism agents) | 5 | 0.55 | 0.23 | 0.60 | 0.40-0.80 | 0.20-1.00 |
| NBT (non-bioterrorism syndromes) | 13 | 0.81 | 0.11 | 0.85 | 0.77-0.92 | 0.54-1.00 |
| Within-participant difference (BT – NBT) | – | -0.26 | 0.21 | -0.22 | -0.45 to -0.12 | -0.72 to +0.03 |

Recognition varied markedly across the five Category A agents, ranging from 24.0% for smallpox to 96.0% for botulism – a more than fourfold difference. Two agents were recognised by the great majority of participants: botulism (24/25, 96.0%, 95% CI [80.5%, 99.3%]) and Ebola virus disease (22/25, 88.0%, 95% CI [70.0%, 95.8%]). The other three were recognised by fewer than half of participants: anthrax (10/25, 40.0%, 95% CI [23.4%, 59.3%]), pneumonic plague (7/25, 28.0%, 95% CI [14.3%, 47.6%]), and smallpox (6/25, 24.0%, 95% CI [11.5%, 43.4%]). The distribution was bimodal: no agent had a recognition rate between 41% and 87%. Per-agent recognition is summarised in Table 6.

**Table 6:** Recognition of each Category A bioterrorism agent (n = 25). Note: Proportion correct is the number of participants correctly identifying the agent from the relevant clinical vignette, divided by the total number of participants (n = 25). 95% binomial confidence intervals were calculated using the Wilson score method without continuity correction.

| Agent | Correct (n/25) | Proportion correct | 95% CI |
| --- | --- | --- | --- |
| Botulism | 24 | 0.96 | 0.81-0.99 |
| Ebola virus disease | 22 | 0.88 | 0.70-0.96 |
| Anthrax | 10 | 0.40 | 0.23-0.59 |
| Pneumonic plague | 7 | 0.28 | 0.14-0.48 |
| Smallpox | 6 | 0.24 | 0.12-0.43 |

## LIMITATIONS

Recruitment was constrained by the absence of MSC approval. Only six of 41 invited schools confirmed dissemination; seven declined predominantly citing absent MSC approval (suggesting MSC routing is now an enforceable expectation for many UK medical schools); and 28 did not reply. Each Head was contacted once with no follow-up.

A definitive response rate could not be calculated. Plausible contributors to low response include survey timing during final-year examination preparation; high baseline survey fatigue; the absence of MSC endorsement potentially weakening perceived legitimacy; and the absence of reminder invitations.

The 25-participant self-selected volunteer sample is unlikely to be representative of UK final-year medical students. At least five schools were identifiable, but the sample is institutionally concentrated relative to the 41 schools with final-year cohorts, and the over-representation of undergraduate-entry participants (24/25) precluded the programme-type comparison. Students confident in their general diagnostic ability may have been more likely to volunteer for an optional diagnostic-vignette survey, though the direction of resulting bias is uncertain given that physician self-assessment correlates poorly with measured competence.^71^

The instrument assesses syndrome recognition but not management – treatment, isolation, public health notification, or broader incident response – so it captures a necessary but insufficient component of appropriate care. The single-best-answer multiple-choice format permits guessing (chance probability 20.0%), so the absolute BT proportion of 0.55 is plausibly an overestimate of knowledge, though this should not systematically bias the within-participant comparison. Tularaemia was not included as its syndrome cannot be reliably distinguished from naturally occurring infections,^25,61^ so the instrument measures only five of the six Category A agents. Each Category A agent was represented by a single vignette, so the per-agent recognition rates should be read as provisional indications rather than stable estimates.

More broadly, written vignettes ask participants how they would manage a hypothetical case in the absence of real clinical context, competing demands, and time pressures, so vignette responses do not necessarily reflect real-world performance.^72–74^

## DISCUSSION

This pilot generated preliminary evidence on Category A syndrome recognition by final-year UK medical students – a previously unstudied population – and tested the feasibility of an instrument and recruitment approach for a subsequent definitive study. The primary finding was that participants performed substantially worse on BT vignettes (M = 0.55) than on NBT vignettes (M = 0.81), a within-participant difference of −0.26 (95% CI [-0.35, −0.18]; t(24) = -6.33, p < 0.001; Cohen’s d_z_ = -1.27), with 24 of 25 participants showing the effect. Per-agent recognition was bimodal: botulism (96.0%) and Ebola virus disease (88.0%) were recognised by the great majority, while anthrax (40.0%), pneumonic plague (28.0%), and smallpox (24.0%) were each recognised by fewer than half. Smallpox was the least well recognised. Three feasibility findings emerged: only six of 41 invited schools confirmed dissemination (seven declined, mostly citing absent MSC approval; 28 did not reply); a definitive response rate could not be calculated; and the planned undergraduate/graduate-entry comparison was infeasible given that only one participant was on a graduate-entry programme.

To the authors’ knowledge, this is the first study to assess UK diagnostic capability for Category A syndromes using tested vignette-based accuracy rather than self-reported confidence; prior UK work has examined curriculum inclusion^68^ but not learner outcomes, and the most directly comparable assessment of tested accuracy^61^ is over two decades old and was conducted at a different career stage in a different country. The instrument has reasonable content validity through iterative expert review by three consultant specialists. The within-participant paired design controls for individual-level variation and yielded a very large effect (Cohen’s d_z_ = -1.27) with only 25 participants, supporting use of the same design in a larger study. The pilot also generated feasibility findings of direct value to subsequent studies.

The within-participant design controls for individual-level variation in baseline diagnostic competence, motivation, and engagement; the same individuals who identified NBT syndromes with high accuracy (mean 0.81) underperformed substantially on BT vignettes. The 26-percentage-point gap therefore reflects a deficit specific to Category A clinical knowledge rather than generally weak diagnostic skills. The finding is broadly consistent with Cosgrove et al.’s 46.8% mean diagnostic accuracy among US clinicians,^61^ though direct comparison is limited by differences in population, instrument, and time period. Other published assessments have relied on self-reported confidence^57,58^ or broader knowledge measures;^59,60^ the present pilot adds UK evidence – in pilot form – that diagnostic underperformance for Category A syndromes appears to persist across geographies, populations, and the two decades since Cosgrove et al.

The bimodal per-agent distribution is striking. The well-recognised agents may benefit from distinctive features, although these explanations are speculative without curriculum data. Botulism’s characteristic descending paralysis with autonomic features and clear exposure history^21^ may aid recognition, while Ebola virus disease has drawn substantial professional attention since the 2014 West African outbreak^75^ and its inclusion within the UK HCID framework.^51^ Smallpox was the least-recognised agent, and this is of particular concern: smallpox is the only Category A agent that no longer occurs naturally, having been declared eradicated by the WHO in 1980.^76^ Any contemporary case must therefore be a laboratory accident or a deliberate release, so failure to recognise smallpox carries a direct biosecurity consequence. The poor recognition is surprising in light of recent mpox outbreaks, including the 2024 WHO-declared Public Health Emergency of International Concern,^77,78^ which might have been expected to raise awareness of poxvirus syndromes including the closely related variola virus.

A plausible explanation for under-recognition is limited curricular coverage: only 17.65% of UK and Irish medical schools reported specific teaching on biological weapons and bioterrorism,^68^ against a broader backdrop of absent terror medicine training in pre-qualification curricula^67^ Clinical exposure during training is concentrated among common presentations, and Category A agents are vanishingly rare in current UK practice, offering little experiential reinforcement. These explanations are provisional and would benefit from formal investigation.

If the pilot findings are confirmed by a definitive study, the implications for UK medical education would be substantial: a specific competence gap that existing UK medical education does not adequately address in a cohort within months of independent NHS practice. Further work would be required to establish the gap’s causes before targeted educational interventions could be designed.

The implications would also extend to UK biosecurity preparedness. Confirmed diagnostic gaps at the point of entry to NHS practice would constitute a substantive weakness in the national capacity to detect a deliberate release, with consequent risks of delayed recognition and response and increased onward transmission, mortality, and social disruption. Addressing this weakness would warrant treatment as a national biosecurity priority, with implications for both medical school curricula and postgraduate training. Further, biosecurity investment has largely moved upstream into pathogen access controls, synthesis screening, and the governance of AI and biological design tools, with comparatively little into the clinical frontline where a release that evaded those controls would first be seen. Strengthening clinical recognition does not compete with those upstream measures.

The pilot findings and feasibility limitations together inform a definitive study to establish the diagnostic capability of final-year UK medical students for Category A syndromes. The validated instrument would be retained but expanded, with several vignettes per Category A agent to yield more stable per-agent estimates, and each participant’s medical school captured. The principal methodological change would be recruitment via the MSC’s Education Leads Advisory Group with prior MSC approval, with timing planned to avoid the final-year examination preparation period and reminders issued through standard MSC procedures. The analyses previously described would be conducted, with the primary paired-samples t-test retained and the programme-type comparison and per-agent recognition analyses undertaken on the assumption of sufficient sample size. The broader limitation of vignette-based assessment would nonetheless persist.

A subsequent study would be required to establish the diagnostic capability of practising UK clinicians – the workforce on which Category A release recognition ultimately depends. The pre-qualification baseline from the definitive student study would provide a useful comparator and inform sample size and analytical design. The expanded instrument developed for the definitive student study would be used, ensuring comparability with the pre-qualification baseline, with recruitment focused on specialties most likely to encounter a Category A presentation – emergency medicine, acute medicine, critical care, and primary care – and cross-specialty comparison a key analytical objective. Recruitment via clinical Royal Colleges would replace the MSC route, and Health Research Authority approval and NHS Research Ethics Committee review would be required.

## CONCLUSION

This pilot study generated preliminary evidence on the diagnostic capability of final-year UK medical students for the syndromes of Category A bioterrorism agents. Participants performed substantially worse on BT vignettes than on NBT vignettes drawn from the same instrument, with a within-participant difference of approximately 26 percentage points, a very large standardised effect size, and a pattern consistent across 24 of 25 participants. Recognition varied markedly across the five agents, with smallpox — whose recognition is diagnostic of a laboratory accident or a deliberate release – the least well recognised. These findings are preliminary and constrained by the pilot recruitment approach, but they motivate a clear research agenda: a definitive student study with MSC approval, followed by a study of practising clinicians in the specialties most likely to encounter a Category A presentation. If confirmed, the implications would extend beyond UK medical education to UK biosecurity preparedness.

## Data Availability

All data produced in the present study are available upon reasonable request to the authors

